# Global and local genetic correlation between congenital heart disease and neurodevelopmental disorders shared genetic architecture

**DOI:** 10.64898/2025.12.13.25342194

**Authors:** Daniel Linares, Beatriz Luna

## Abstract

Individuals with congenital heart disease (CHD) have an increased risk of neurodevelopmental disorders (NDDs), a relationship likely driven by multiple interacting factors, including shared genetic influences, the underlying cardiac malformation, and risks associated with medical and surgical care. To investigate the genetic component of this association, we analyzed genome-wide association study (GWAS) summary statistics for CHD from the UK Biobank and FinnGen, focusing on two phenotypes present in both cohorts: congenital malformations of cardiac septa (Q21) and congenital malformations of great arteries (Q25). A fixed-effect meta-analysis was performed using METAL to increase statistical power. Neurodevelopmental phenotypes were represented by Austism Spectrum Disorder (ASD) and Attention-deficit/hyperactivity disorder (ADHD) GWAS results from the Psychiatric Genomics Consortium. Genome-wide genetic correlations were estimated using the High-Definition Likelihood. Local genetic correlations were assessed using LAVA. Multiple testing was controlled using Bonferroni correction.

No significant genome-wide genetic correlations were observed between CHD phenotypes and either ASD or ADHD. However, LAVA identified a single significant local genetic correlation between cardiac septal defects and ASD on chromosome 6. Within this block, two loci near *GMDS* showed significant association with cardiac septal malformations. Bayesian colocalization did not reveal a variant with strong evidence for a shared causal signal, though three *GMDS* variants demonstrated moderate support for colocalization.

These findings suggest that while global genetic overlap between CHD and NDDs is limited, specific loci, such as the *GMDS* region on chromosome 6, may contribute to shared developmental pathways underlying both phenotypes.

## Introduction

Congenital heart disease (CHD) is the most prevalent congenital anomaly, affecting approximately 1% of live births globally (1). Advances in medical and surgical management have significantly improved survival, shifting attention toward long-term neurodevelopmental outcomes in this population (1,2). Numerous epidemiological studies have demonstrated that children with CHD, particularly those with complex lesions, are at increased risk for a broad spectrum of neurodevelopmental disorders (NDD), including autism spectrum disorder (ASD), attention-deficit/hyperactivity disorder (ADHD), intellectual disability, and speech and language impairments (3–6). These cognitive impairments persist throughout adolescence and adulthood.

The impact of CHD on neurodevelopment is likely the result of multiple interacting factors, including genetic influences, the underlying heart defect itself, and risks associated with medical and surgical treatment. In each individual, these factors likely combine and reinforce one another, beginning as early as fetal development and continuing throughout childhood (7).

Despite this strong epidemiological evidence, the extent to which CHD and NDD share a common genetic basis remains unclear. Syndromic forms of CHD, such as 22q11.2 deletion syndrome and CHARGE syndrome, support a shared genetic etiology, as they often present with both heart defects and neurodevelopmental impairments in their phenotype (8). In addition, studies have shown that *de novo* mutations affecting key developmental pathways and structural variants may contribute to both CHD and NDD (9), providing further support for a shared etiology.

The aim of this study was to investigate the shared genetic architecture between congenital heart defects and neurodevelopmental disorders, using both genome-wide and local genetic correlation. We sought to identify specific loci that contribute to risk in both phenotypes and to evaluate whether any signals reflect shared causal variants. Through this framework, we further aimed to characterize the biological relevance of implicated genes and pathways, providing insight into potential mechanisms underlying the co-occurrence of cardiac and neurodevelopmental anomalies.

## Methods

### Data sources and variant‐level quality control

Genome-wide association study (GWAS) summary statistics for congenital heart disease (CHD) were obtained from the UK Biobank (UKBB) (10), FinnGen release 12 (11). We selected two CHD phenotypes that were present in both datasets:

- Q21 – Congenital malformations of cardiac septa
- Q25 – Congenital malformations of great arteries

To increase statistical power and improve reliability, we conducted a fixed-effect meta-analysis across both cohorts using the software METAL, applying the sample-size–based method to combine effect estimates across studies.

For **neurodevelopmental disorders (NDDs)**, we used GWAS summary statistics for ASD (12) and ADHD (13) from the Psychiatric Genomics Consortium (PGC). The total sample sizes and case–control numbers for each dataset are provided in Table 1.

**Table 1.** Genome-wide association studies included in the study.

| Consortium | Phenotype | Sample size |
| --- | --- | --- |
| FinnGen r12 | Congenital malformations of great arteries | 687 / 494430 |
| FinnGen r12 | Septal defects | 2184 / 497928 |
| UK BioBank | Q25 Congenital malformations of great arteries | 111 / 420420 |
| UK BioBank | Q21 Congenital malformations of cardiac septa | 687 / 419844 |
| Psychiatric Genomic Consortium | Autism spectrum disorder | 18382 / 27969 |
| Psychiatric Genomic Consortium | Attention-deficit/hyperactivity disorder | 38691 / 186843 |

We included only **single nucleotide polymorphisms (SNPs)** that were present in all three datasets (UKBB, FinnGen, and PGC). Analyses were restricted to autosomal single nucleotide polymorphisms (SNPs) present in the HapMap-3 panel, excluding the major histocompatibility complex (MHC) region, and required minor allele frequency (MAF) > 0.01. Variants with reference/alternate allele mismatches relative to the reference panel were removed prior to analysis.

### Global genetic correlation

We estimated genome-wide genetic correlations (r_g_) between CHD and each neurodevelopmental phenotype using the High-Definition Likelihood (HDL) framework. HDL provides a dense pre-computed LD reference panel (1,029,876 quality-controlled HapMap-3 SNPs from UK Biobank). Compared to LD Score regression (LDSC), HDL reduces the variance of a genetic correlation estimate by about 60% (14). We applied HDL using the provided precomputed reference LD matrices, restricting the analysis to variants that passed quality control: common SNPs (MAF > 0.01), present in HapMap3, outside the MHC region, and with no reference allele mismatches.

### Local genetic correlation

To localize shared heritability, we used the LAVA (Local Analysis of [co]Variant Association) method (15). We used the linkage disequilibrium (LD) reference matrices from 1000 Genomes Phase 3 European samples, using chromosome‐position coordinates in human genome build 37 and PLINK format, provided by MAGMA (https://cncr.nl/research/magma/).

Each GWAS summary was partitioned into 2,495 largely independent LD blocks as per the original LAVA preprint. For every LAVA analysis we selected blocks (loci) that had significant hits from at least one trait, as suggested by Reynolds R. and collaborators (16). Then, we performed a univariate heritability (h^2^) test within each block, and only blocks with significant univariate h^2^ (p < 0.05, Bonferroni‐corrected) in both traits underwent bivariate local r_g_ estimation. We controlled for multiple testing across all bivariate comparisons by applying the Bonferroni method.

### Colocalization of shared loci

For each locus with significant local genetic correlation, we performed Bayesian colocalization analysis using the coloc.abf algorithm. Posterior probability for a shared causal variant (PP4) ≥ 0.8 was taken as strong evidence for colocalization. This step aim to yield a set of implicated SNPs at each locus.

### Gene mapping

Implicated SNPs from colocalization were mapped to the nearest gene by physical proximity. We queried Ensembl via the biomaRt interface to identify the closest protein‐coding gene to each lead SNP.

## Results

The genome-wide genetic correlation between CHD and NDD from HDL framework revealed no significant correlation (Table 2).

**Table 2.** Genome-wide and local genetic correlation of CHD on NDD.

| Genomic Region | Phenotype with GWAS hits in locus | $r_g$ | 95% CI | p | p (adjusted) |
| --- | --- | --- | --- | --- | --- |
| Autism Spectrum Disorder - Congenital malformations of cardiac septa |  |  |  |  |  |
| Genome-wide (HDL) | n/a |  |  |  |  |
| 6:1380680-2746531 (*GMDS*) | Q21 / ASD<br>$1.42 \times 10^{-8} / 3.34 \times 10^{-4}$ | -0.667 | -1/-0.297 | **0.00078** | **0.0186** |
| 10:72659224-73514214 | Q21 / ASD<br>$2.17 \times 10^{-10} / 9.14 \times 10^{-6}$ | 0.181 | -0.161/0.548 | 0.283 | 1 |
| 18:55809316-56832312 | Q21 / ASD<br>$1.9 \times 10^{-4} / 3.01 \times 10^{-5}$ | 0.0512 | -0.418/0.554 | 0.818 | 1 |
| Attention-Deficit/Hyperactivity Disorder - Congenital malformations of cardiac septa |  |  |  |  |  |
| Genome-wide (HDL) | n/a |  |  |  |  |
| 1:69438910-70992905 | Q21 / ADHD<br>$7.05 \times 10^{-5} / 1.58 \times 10^{-4}$ | -0.460 | -1/-0.0187 | 0.044 | 0.6162 |
| 2:10611655-11230349 | Q21 / ADHD<br>$1.52 \times 10^{-5} / 2.68 \times 10^{-9}$ | -0.437 | -0.856/-0.089 | 0.015 | 0.2142 |
| 2:215899571-217566011 | Q21 / ADHD<br>$1.44 \times 10^{-4} / 9.6 \times 10^{-6}$ | 0.125 | -0.314/0.593 | 0.556 | 1 |
| 8:92876621-94999066 | Q21 / ADHD<br>$5.53 \times 10^{-5} / 2.8 \times 10^{-19}$ | -0.166 | -0.498/0.140 | 0.283 | 1 |
| 11:133521643-134351064 | Q21 / ADHD<br>$8.35 \times 10^{-5} / 1.41 \times 10^{-9}$ | -0.0847 | -0.485/0.298 | 0.644 | 1 |

| Autism Spectrum Disorder - Congenital malformations of great arteries |  |  |  |  |  |
| --- | --- | --- | --- | --- | --- |
| Genome-wide (HDL) | n/a |  |  |  |  |
| 5:113121556-113998532 | Q25 / ASD<br>5.84*10<sup>-4</sup>/3.32*10<sup>-4</sup> | 0.457 | -0.0514/1 | 0.072 | 1 |
| Attention-Deficit/Hyperactivity Disorder - Congenital malformations of great arteries |  |  |  |  |  |
| Genome-wide (HDL) | n/a |  |  |  |  |
| 2:10611655-11230349 | Q25 / ADHD<br>1.53*10<sup>-6</sup>/3.02*10<sup>-9</sup> | 0.0334 | -0.318/0.376 | 0.845 | 1 |
| 9:85135752-86769886 | Q25 / ADHD<br>4.07*10<sup>-11</sup>/1.39*10<sup>-6</sup> | -0.312 | -0.656/0.0026 | 0.0495 | 0.6927 |
| 9:121008530-122677719 | Q25 / ADHD<br>6.05*10<sup>-10</sup>/6.37*10<sup>-5</sup> | 0.0514 | -0.343/0.443 | 0.778 | 1 |
| 17:1480057-2699135 | Q25 / ADHD<br>3.83*10<sup>-5</sup>/3.22*10<sup>-10</sup> | -0.253 | -0.658/0.092 | 0.157 | 1 |
| 18:5575813-6756496 | Q25 / ADHD<br>2.31*10<sup>-4</sup>/3.37*10<sup>-6</sup> | -0.0765 | -0.545/0.380 | 0.719 | 1 |

We identify the flowing number for LD blocks harboring signals from each of the trait peers:

- Congenital malformations of cardiac septa with ADHD = 175 blocks, 5 of them showed significant univariate heritability in either trait.
- Congenital malformations of great arteries with ADHD = 166 blocks, 5 of them showed significant univariate heritability in either trait.
- Congenital malformations of cardiac septa with ASD = 56 blocks, 3 of them showed significant univariate heritability in either trait.
- Congenital malformations of great arteries with ASD = 63 blocks, 1 of them showed significant univariate heritability in either trait.

LAVA identified one significant local genetic correlation between congenital malformations of cardiac septa and ASD in chromosome 6 (Table 2 and Figure 1). Inside this block, two loci near *GMDS* had significant hits for cardiac septa malformations only.

**Figure 1.**
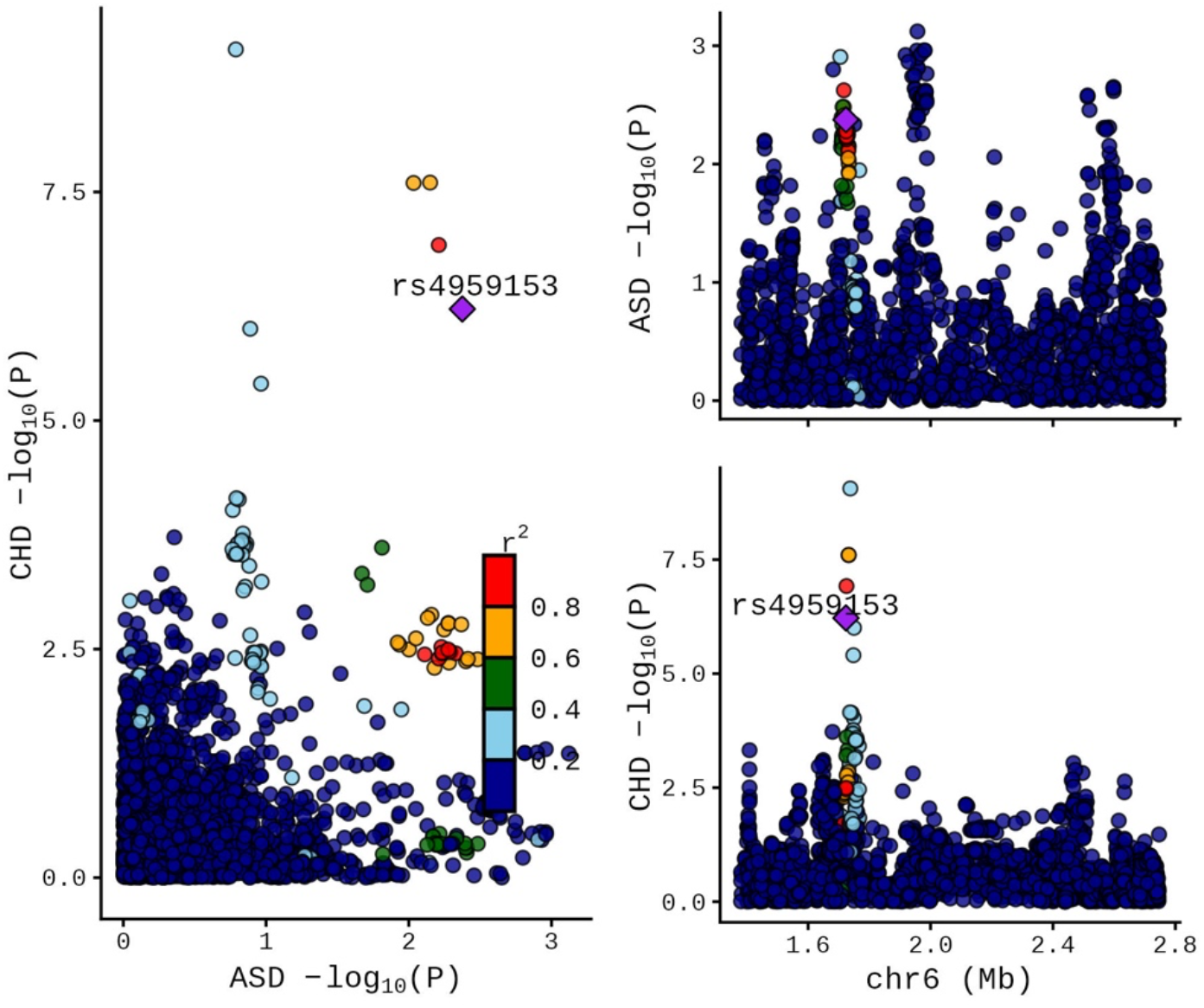
LocusCompare plots of LAVA results on Austism Spectrum Disorders and Congenital malformations of cardiac septa.

Bayesian colocalization analysis of the chromosome 6 locus did not identify any single variant with a high posterior probability of representing a shared causal signal. Nonetheless, three variants within *GMDS* showed moderate evidence of colocalization: rs61207262 (PP_4_ = 0.398), rs1986205 (PP_4_ = 0.283), and rs1986204 (PP_4_ = 0.221)

## Discussion

In this study, we examined the shared genetic architecture between congenital heart defects (CHD) and neurodevelopmental disorders (NDD) using both genome-wide and local correlation. Genome-wide genetic correlation analyses using the HDL method revealed no significant correlation between CHD subtypes and NDD. Local genetic correlation, using LAVA, detected a single significant local genetic correlation block between congenital malformations of cardiac septa and ASD on chromosome 6. Within this region, two loci near *GMDS* displayed significant association with cardiac septal defects.

Observational studies have consistently shown that children with CHD are at increased risk of neurodevelopmental and behavioral problems (3). Children with CHD exhibited an increased odds of anxiety and depression, among other adverse psychosocial outcomes (17,18). For instance, in the Boston Circulatory Arrest Study those with transposition of the great vessels had a five-to seven-fold increased likelihood of ADHD diagnosis (19). A meta-analysis by Gu et al. similarly found that CHD approximately doubled ASD risk (20). Liu et al. analyzed 85 314 participants and showed associations between CHD and intellectual disability, speech and language disorders, and ADHD, with CHD severity correlating with ASD prevalence (4). More recently, Miller et al. found that in early adolescence, transplant-free survivors of surgical palliation for hypoplastic left heart syndrome show concerning impairments across all domains of neurodevelopment, including intelligence, attention, executive functioning, social development, visual memory, and adaptive functioning (21).

The low incidence rate of CHD imposes practical constraints, as most studies have small sample sizes and often focus on complex, syndromic lesions such as hypoplastic left heart syndrome (6,21,22). While these severe phenotypes may carry a stronger mendelian component (23,24), they also entail more extensive cyanosis, surgical interventions, and perioperative complications, all of which can independently affect neurodevelopment (3,7). Thus, the interplay of genetic and non-genetic factors in CHD-associated neurobehavioral outcomes is particularly challenging to disentangle.

There is, however, clear evidence for a shared genetic etiology of CHD and NDD. Syndromic forms of CHD, such as CHARGE syndrome and 22q11.2 deletion syndrome, commonly include ASD and other neurodevelopmental phenotypes (8). A review by Vorstman and colleagues identified at least 10 chromosomal anomalies co-occurring with both CHD and ASD (25). Furthermore, damaging *de novo* mutations in genes linked to autism have been reported in children with CHD, and these may disrupt key developmental pathways (9,26,27). Both the heart and brain derive, in part, from neural crest cells and share overlapping morphogenetic pathways, including critical transcriptional regulators and chromatin remodeling complexes (26). Common variants could also play a role in the shared etiology of CHD and NDD, polymorphisms of APOE have been implicated in impaired neurodevelopmental outcomes in congenital heart disease (28).

In addition, it is now well established that the genetic basis of ASD is dominated by rare, high-penetrance de novo variants, although polygenic risk does contribute to overall liability (29–31). ADHD, by contrast, shows stronger polygenic architecture, with common variants explaining a larger proportion of its heritability, albeit each contributing very small effects (32). CHD, particularly in its non-syndromic form, is genetically heterogeneous and likely includes both polygenic and oligogenic components, but also shows consistent association with environmental exposures (23,33).

*GMDS* locus, one of the regions highlighted by LAVA, has not been previously identified as a potential risk factor for ASD. *GMDS* encodes GDP-mannose 4,6-dehydratase, the enzyme that catalyzes the first step in the conversion of GDP-mannose to GDP-fucose, initiating the fucosylation pathway (34). Fucosylation is essential for several processes relevant to development (35). *GMDS* does not show strong evidence of intolerance to loss-of-function variants (pLI = 0.54), its missense Z-score (0.5) suggests no significant selective pressure against missense variants (36), and, given that the GMDS variants showing colocalization are common in the non-Finnish European population (MAF ∼0.2), these alleles are likely to be benign. Consequently, any contribution of *GMDS* to CHD or ASD risk is likely to reflect a more complex genetic architecture driven by common polymorphisms rather than highly penetrant rare variants. This interpretation is consistent with the reliance on GWAS data, which predominantly captures the effects of common variation and is less suited to detecting rare, high-impact mutations.

Structural variants (deletions) involving the *GMDS* locus has been associated with both neurodevelopmental and cardiac defects, including classic or mild Dandy–Walker malformation, mega cisterna magna, cerebellar vermis hypoplasia (37), Ebstein anomaly, and tetralogy of Fallot (35). These phenotypes point toward a role for disrupted fucosylation in the development of both the central nervous system and the heart.

The local genetic correlation observed between ASD and congenital heart defects at this locus may therefore reflect shared developmental pathways. There is some experimental evidence suggesting that *GMDS* interacts functionally with components of the Notch signaling pathway (38), a highly conserved mechanism involved in intercellular communication between multiple organ systems, cell development, and differentiation (39). Notch signaling is particularly active during early cortical neurogenesis and cardiac morphogenesis (39). Moreover, of the 69 gene variants associated with both CHD and NDD reported by Homsy et al., 32 involved transcriptional regulators, including components of the Wnt and Notch signaling pathways (26).

Although *GMDS* has not been previously linked to ASD, its biological function and the phenotypes related to this gene provide plausible mechanistic routes through which variation in this region could contribute to risk in both conditions. Further functional validation and fine-mapping will be needed to determine whether the signal is driven by *GMDS* itself, regulatory elements in its vicinity, or neighboring genes within the locus.

Beyond genetics, CHD-associated neurodevelopmental risk is amplified by environmental insults inherent to treating congenital defects (7). Cardiopulmonary bypass in infancy and chronic cyanosis impose hypoxic and inflammatory stress on the developing brain (3,40,41). Fetal neurimaging studies have found that brain dysmaturation, in cases with CHD, begins in utero and may linked to altered cerebral blood flow (42,43). Other contributors include prematurity, feeding delays, parental psychological distress, and socioeconomic disadvantages (3). Recent studies suggest that placental dysregulation may contribute to altered fetal brain and cardiac development and explain at least part of CHD-associated NDD (44). None of these factors are mutually exclusive, so more than one may contribute to the relationship between CHD and NDD in a given individual.

Epigenetic mechanisms have received comparatively little attention in studies of the shared etiology between CHD and NDD, despite growing evidence that genetic mutations account for only a minority of CHD cases. Aberrant gene expression driven by epigenetic dysregulation appears to play a major role in the pathogenesis of CHD (45), and similar mechanisms are increasingly implicated in neurodevelopment. Multiple epigenetic processes, including DNA methylation, histone modification, and noncoding RNAs, regulate key aspects of neuronal differentiation, and disruptions of these marks have been linked to a range of NDD phenotypes (46). Likewise, mutations in genes that control chromatin structure, such as CHD7, are known to impair both heart and brain development, exemplified by CHARGE syndrome (28). Furthermore, epigenetic regulation within the placenta can influence fetal growth by altering placental function and signaling pathways; notably, trophoblast DNA methylation patterns have been strongly associated with CHD (44). Together, these findings highlight epigenetic regulation as a plausible yet underexplored mechanism underlying the co-occurrence of CHD and NDD.

Several limitations should be acknowledged when interpreting these findings. First, although we increased sample size for the CHD phenotypes through meta-analysis using METAL, the absolute number of CHD cases remained modest, which limited statistical power to detect smaller effects or additional loci with shared genetic architecture. Second, discrepancies in SNP coverage across datasets constrained the scope of the local genetic correlation and colocalization analyses. FinnGen and UK Biobank included approximately 21 million and 85 million variants, respectively, whereas the PGC ASD and ADHD GWAS summary datasets contained only ∼6 million SNPs. As a result, many variants, particularly those surrounding GMDS that were strongly associated with cardiac septal defects in FinnGen dataset, were absent from the PGC datasets and therefore excluded from the analysis. This mismatch may have reduced our ability to detect shared causal variants. Finally, all analyses were restricted to individuals of European ancestry. The generalizability of these results to other ancestries remains uncertain, and replication in more diverse cohorts will be essential to determine the broader relevance of the identified signals.

## Conclusion

In summary, our study revealed no evidence of genome-wide genetic correlation between congenital heart defects and neurodevelopmental disorders yet identified a significant local genetic overlap between cardiac septal defects and ASD at a locus on chromosome 6. Signals within this region pointed toward GMDS, a gene with biologically plausible roles in both cardiac and neural development. These findings highlight a potentially novel locus linking CHD and NDD. Further studies in larger cohorts will be essential to validate this association and clarify the functional contribution of GMDS to early developmental pathways.

## Data availability

The Genome Wide Association Study (GWAS) summary statistics used in this study are public and can be accessed via FinnGen database (https://www.finngen.fi/en/access_results), UK Biobank (https://pan.ukbb.broadinstitute.org) and the PGC database (https://pgc.unc.edu/for-researchers/download-results/).

## Code availability

All codes related to this manuscript are available on Github repository: https://github.com/Dani12alt/genetic_correlation_chd_ndd.git

## Acknowledgements

We want to thank Dr. Maria Soler Artigas from the Vall d’Hebron Institut de Recerca, Barcelona, Spain. Without her valuable insights this study would not be possible.

Also, we want to acknowledge the participants and investigators of the FinnGen study, UK Biobank, and PGC. This article would have been hard to complete without the support and commitment of these organizations and their members.

## References

1. Wu W, He J, Shao X. Incidence and mortality trend of congenital heart disease at the global, regional, and national level, 1990–2017. Medicine. 2020;99(23):e20593. 10.1097/MD.0000000000020593.

2. Zimmerman MS, Smith AGC, Sable CA, Echko MM, Wilner LB, Olsen HE, et al. Global, regional, and national burden of congenital heart disease, 1990–2017:a systematic analysis for the Global Burden of Disease Study 2017. The Lancet Child & Adolescent Health. 2020;4(3):185–200. 10.1016/S2352-4642(19)30402-X.

3. Sood E, Newburger JW, Anixt JS, Cassidy AR, Jackson JL, Jonas RA, et al. Neurodevelopmental Outcomes for Individuals With Congenital Heart Disease: Updates in Neuroprotection, Risk-Stratification, Evaluation, and Management: A Scientific Statement From the American Heart Association. Circulation. 2024;149(13):e997–e1022. 10.1161/CIR.0000000000001211.

4. Liu Z yuan, Wang Q qiong, Pang X yong, Huang X bi, Yang G ming, Zhao S. Association of congenital heart disease and neurodevelopmental disorders: an observational and Mendelian randomization study. Italian Journal of Pediatrics. 2024;50(1):63. 10.1186/s13052-024-01610-3.

5. Tsao PC, Lee YS, Jeng MJ, Hsu JW, Huang KL, Tsai SJ, et al. Additive effect of congenital heart disease and early developmental disorders on attention-deficit/hyperactivity disorder and autism spectrum disorder: a nationwide population-based longitudinal study. European Child & Adolescent Psychiatry. 2017;26(11):1351–1359. 10.1007/s00787-017-0989-8.

6. Newburger JW, Sleeper LA, Bellinger DC, Goldberg CS, Tabbutt S, Lu M, et al. Early Developmental Outcome in Children With Hypoplastic Left Heart Syndrome and Related Anomalies. Circulation. 2012;125(17):2081–2091. 10.1161/CIRCULATIONAHA.111.064113.

7. Howell HB, Zaccario M, Kazmi SH, Desai P, Sklamberg FE, Mally P. Neurodevelopmental outcomes of children with congenital heart disease: A review. Current Problems in Pediatric and Adolescent Health Care. 2019;49(10):100685. 10.1016/j.cppeds.2019.100685.

8. Richards C, Jones C, Groves L, Moss J, Oliver C. Prevalence of autism spectrum disorder phenomenology in genetic disorders: a systematic review and meta-analysis. The Lancet Psychiatry. 2015;2(10):909–916. 10.1016/S2215-0366(15)00376-4.

9. Wang YJ, Zhang X, Lam CK, Guo H, Wang C, Zhang S, et al. Systems analysis of de novo mutations in congenital heart diseases identified a protein network in the hypoplastic left heart syndrome. Cell Systems. 2022;13(11):895-910.e4. 10.1016/j.cels.2022.09.001.

10. UK Biobank Whole-Genome Sequencing Consortium. Whole-genome sequencing of 490,640 UK Biobank participants. Nature. 2025;645(8081):692–701. 10.1038/s41586-025-09272-9.

11. Kurki MI, Karjalainen J, Palta P, Sipilä TP, Kristiansson K, Donner KM, et al. FinnGen provides genetic insights from a well-phenotyped isolated population. Nature. 2023;613(7944):508–518. 10.1038/s41586-022-05473-8.

12. Grove J, Ripke S, Als TD, Mattheisen M, Walters RK, Won H, et al. Identification of common genetic risk variants for autism spectrum disorder. Nature Genetics. 2019;51(3):431–444. 10.1038/s41588-019-0344-8.

13. Demontis D, Walters GB, Athanasiadis G, Walters R, Therrien K, Nielsen TT, et al. Genome-wide analyses of ADHD identify 27 risk loci, refine the genetic architecture and implicate several cognitive domains. Nature Genetics. 2023;55(2):198–208. 10.1038/s41588-022-01285-8.

14. Ning Z, Pawitan Y, Shen X. High-definition likelihood inference of genetic correlations across human complex traits. Nature Genetics. 2020;52(8):859–864. 10.1038/s41588-020-0653-y.

15. Werme J, van der Sluis S, Posthuma D, de Leeuw CA. An integrated framework for local genetic correlation analysis. Nature Genetics. 2022;54(3):274–282. 10.1038/s41588-022-01017-y.

16. Reynolds RH, Wagen AZ, Lona-Durazo F, Scholz SW, Shoai M, Hardy J, et al. Local genetic correlations exist among neurodegenerative and neuropsychiatric diseases. npj Parkinson’s Disease. 2023;9(1):70. 10.1038/s41531-023-00504-1.

17. Abda A, Bolduc ME, Tsimicalis A, Rennick J, Vatcher D, Brossard-Racine M. Psychosocial Outcomes of Children and Adolescents With Severe Congenital Heart Defect: A Systematic Review and Meta-Analysis. Journal of Pediatric Psychology. 2019;44(4):463–477. 10.1093/jpepsy/jsy085.

18. Gonzalez VJ, Kimbro RT, Cutitta KE, Shabosky JC, Bilal MF, Penny DJ, et al. Mental Health Disorders in Children With Congenital Heart Disease. Pediatrics. 2021;147(2):e20201693. 10.1542/peds.2020-1693.

19. DeMaso DR, Labella M, Taylor GA, Forbes PW, Stopp C, Bellinger DC, et al. Psychiatric disorders and function in adolescents with d-transposition of the great arteries. The Journal of Pediatrics. 2014;165(4):760–766. 10.1016/j.jpeds.2014.06.029.

20. Gu S, Katyal A, Zhang Q, Chung W, Franciosi S, Sanatani S. The Association Between Congenital Heart Disease and Autism Spectrum Disorder: A Systematic Review and Meta-Analysis. Pediatric Cardiology. 2023;44(5):1092–1107. 10.1007/s00246-023-03146-5.

21. Miller TA, Sharma B, Gongwer R, Trachtenberg FL, Newburger JW, Goldberg CS, et al. Neurodevelopmental Outcomes in Early Adolescence: The Pediatric Heart Network’s Single Ventricle Reconstruction Trial. Circulation. 2025;152(17):1246–1261. 10.1161/CIRCULATIONAHA.125.074523.

22. Ricci MF, Andersen JC, Joffe AR, Watt MJ, Moez EK, Dinu IA, et al. Chronic Neuromotor Disability After Complex Cardiac Surgery in Early Life. Pediatrics. 2015;136(4):e922–933. 10.1542/peds.2015-1879.

23. Blue GM, Kirk EP, Giannoulatou E, Sholler GF, Dunwoodie SL, Harvey RP, et al. Advances in the Genetics of Congenital Heart Disease: A Clinician’s Guide. Journal of the American College of Cardiology. 2017;69(7):859–870. 10.1016/j.jacc.2016.11.060.

24. Grossfeld P, Ye M, Harvey R. Hypoplastic Left Heart Syndrome. JACC. 2009;53(12):1072– 1074. 10.1016/j.jacc.2008.12.024.

25. Vorstman JAS, Parr JR, Moreno-De-Luca D, Anney RJL, Nurnberger JI, Hallmayer JF. Autism genetics: opportunities and challenges for clinical translation. Nature Reviews. Genetics. 2017;18(6):362–376. 10.1038/nrg.2017.4.

26. Homsy J, Zaidi S, Shen Y, Ware JS, Samocha KE, Karczewski KJ, et al. De novo mutations in congenital heart disease with neurodevelopmental and other congenital anomalies. Science. 2015;350(6265):1262–1266. 10.1126/science.aac9396.

27. Jin SC, Homsy J, Zaidi S, Lu Q, Morton S, DePalma SR, et al. Contribution of rare inherited and de novo variants in 2,871 congenital heart disease probands. Nature Genetics. 2017;49(11):1593–1601. 10.1038/ng.3970.

28. Patt E, Singhania A, Roberts AE, Morton SU. The Genetics of Neurodevelopment in Congenital Heart Disease. The Canadian Journal of Cardiology. 2023;39(2):97–114. 10.1016/j.cjca.2022.09.026.

29. Yasuda Y, Matsumoto J, Miura K, Hasegawa N, Hashimoto R. Genetics of autism spectrum disorders and future direction. Journal of Human Genetics. 2023;68(3):193–197. 10.1038/s10038-022-01076-3.

30. Sandin S, Lichtenstein P, Kuja-Halkola R, Hultman C, Larsson H, Reichenberg A. The Heritability of Autism Spectrum Disorder. JAMA. 2017;318(12):1182–1184. 10.1001/jama.2017.12141.

31. Klei L, McClain LL, Mahjani B, Panayidou K, De Rubeis S, Grahnat ACS, et al. How rare and common risk variation jointly affect liability for autism spectrum disorder. Molecular Autism. 2021;12(1):66. 10.1186/s13229-021-00466-2.

32. Faraone SV, Larsson H. Genetics of attention deficit hyperactivity disorder. Molecular Psychiatry. 2019;24(4):562–575. 10.1038/s41380-018-0070-0.

33. Gao H, Liu Y, Sheng W, Shou W, Huang G. Progresses in genetic testing in congenital heart disease. Medicine Plus. 2024;1(2):100028. 10.1016/j.medp.2024.100028.

34. PubChem. GMDS - GDP-mannose 4,6-dehydratase (human). https://pubchem.ncbi.nlm.nih.gov/gene/GMDS/human [Accessed 7th December 2025].

35. Lo-A-Njoe SM, Verberne EA, van der Veken LT, Arends E, van Tintelen JP, Postma AV, et al. GMDS Intragenic Deletions Associate with Congenital Heart Disease including Ebstein Anomaly. Cardiogenetics. 2023;13(3):106–112. 10.3390/cardiogenetics13030010.

36. Chen S, Francioli LC, Goodrich JK, Collins RL, Kanai M, Wang Q, et al. A genomic mutational constraint map using variation in 76,156 human genomes. Nature. 2024;625(7993):92–100. 10.1038/s41586-023-06045-0.

37. Aldinger KA, Lehmann OJ, Hudgins L, Chizhikov VV, Bassuk AG, Ades LC, et al. FOXC1 is required for normal cerebellar development and is a major contributor to chromosome 6p25.3 Dandy-Walker malformation. Nature Genetics. 2009;41(9):1037–1042. 10.1038/ng.422.

38. Ameen MT, Fowler G, French CR. Mutation of the GDP-Fucose Biosynthesis Gene gmds Increases Hair Cell Number and Neuromast Regenerative Capacity in Zebrafish. International Journal of Molecular Sciences. 2025;26(19):9737. 10.3390/ijms26199737.

39. Sajid A, Chavez-Valdez R, Sharp AN, Shah DK. Neurodevelopment in congenital heart disease: a review of antenatal mechanisms and therapeutic potentials. Pediatric Research. 2025; 1–8. 10.1038/s41390-025-04360-y.

40. Zhu S, Sai X, Lin J, Deng G, Zhao M, Nasser MI, et al. Mechanisms of perioperative brain damage in children with congenital heart disease. Biomedicine & Pharmacotherapy. 2020;132:110957. 10.1016/j.biopha.2020.110957.

41. Dhari Z, Leonetti C, Lin S, Prince A, Howick J, Zurakowski D, et al. Impact of Cardiopulmonary Bypass on Neurogenesis and Cortical Maturation. Annals of neurology. 2021;90(6):913–926. 10.1002/ana.26235.

42. Feldmann M, Guo T, Miller SP, Knirsch W, Kottke R, Hagmann C, et al. Delayed maturation of the structural brain connectome in neonates with congenital heart disease. Brain Communications. 2020;2(2):fcaa209. 10.1093/braincomms/fcaa209.

43. Leonetti C, Back SA, Gallo V, Ishibashi N. Cortical Dysmaturation in Congenital Heart Disease. Trends in neurosciences. 2019;42(3):192–204. 10.1016/j.tins.2018.12.003.

44. Demonceaux M, Werner O, Cadeau O, Guerra A, Roy A, Ferchaud-Roucher V, et al. Congenital Heart Diseases and Neurodevelopmental Disorders: New Insights Through the DOHaD Hypothesis. JACC: Basic to Translational Science. 2025;10(8):101251. 10.1016/j.jacbts.2025.01.022.

45. Wang G, Wang B, Yang P. Epigenetics in Congenital Heart Disease. Journal of the American Heart Association. 2022;11(7):e025163. 10.1161/JAHA.121.025163.

46. Reichard J, Zimmer-Bensch G. The Epigenome in Neurodevelopmental Disorders. Frontiers in Neuroscience. 2021;15. 10.3389/fnins.2021.776809.

